# Visual Field Progression Rate and Laminar Depth and Curvature in the African Descent and Glaucoma Evaluation Study (ADAGES)

**DOI:** 10.1101/2024.11.02.24316193

**Authors:** Karla Murillo, Anuwat Jiravarnsirikul, Evan Walker, Massimo A. Fazio, Stuart Gardiner, Robert N Weinreb, Jeffrey M Liebmann, Linda M. Zangwill, Christopher A. Girkin

**Author notes:** **Correspondence** Massimo A. Fazio, Ph.D., Department of Ophthalmology and Visual Sciences, 1720 University Blvd, Birmingham, AL, United States, 35294.

## Abstract

**Purpose:** To determine if lamina cribrosa (LC) depth and curvature predict visual field (VF) progression rate in the African Descent and Glaucoma Evaluation Study (ADAGES) cohort in patients with primary open angle glaucoma (POAG).

**Subjects:** Participants, and/or Controls. Three eyes from three research-consented brain-dead organ donors.

**Methods:** Anterior laminar cribrosa surface depth (ALCSD) and LC curvature index (LCCI) were defined from 24 radial B-scan spectral domain optic coherence tomographic (SDOCT) images of each image. All scans were processed using a deep learning software to perform the segmentation and optic nerve health (ONH) layers. Univariable and multivariable linear mixed effects models were used to test associations between VF progression rate, ALCSD, and other demographic and clinical characteristics and known risk factors for progressive POAG. Additionally, the variable selection in the final multivariable model was reached using the Akaike information criterion index (AICc) and clinical utility.

**Measures:** IOP change with exposure to NP.

**Main Outcome:** The associations between ALCSD and LCCI with the rate of VF progression.

**Results:** There was a statistically significant relationship between faster VF progression rates, and a deeper ALCSD (−0.02 dB/year/50 microns, p<0.001) and LCCI (−0.01unit/dB/year, p<0.001). There was also increasing progression seen in both models for with increasing age (−0.025dB/year/10 years, p<0.007), intraocular pressure (IOP) (−0.011dB/year per 2 mmHg, p=0.021), and disease severity (0.017dB/year per dB, p<0.001). The rate of progression was faster in the ED cohort (−0.056 dB/year, p=0.006) following propensity matching for disease severity and age.

**Conclusions:** In the ADAGES cohort, there is a statistically significant association between VF progression rate and ALCSD and curvature (LCCI). A 50 um deeper LC at baseline had a similar effect of VF progression as being 10 years of age older. These data suggest that morphologic change in the ONH due to glaucomatous and age-related remodeling may induce greater vulnerability to develop progressive disease. Thus, LC depth and curvature may inform the likelihood and rate of glaucoma progression and are promising candidates for mechanistic biomarkers.

## Introduction

The number of people in the United States (US) with glaucoma is expected to increase from 2.7 million to 6.3 million by 2030^1-3^. Glaucoma is a progressive optic neuropathy resulting in loss of retinal ganglion cells (RGC)^4^. A preponderance of evidence suggests that the initial site of injury lies within the optic nerve head (ONH)^5^, a biomechanically dynamic region in which the RGC axons traverse from the relatively high-pressure intraocular compartment to the lower pressure space within the retroorbital optic nerve sheath. The lamina cribrosa (LC) provides mechanical and vascular support to the RGC axons within the ONH. Remodeling of the LC is a histological hallmark of glaucomatous optic neuropathy. Thus, both age-related and glaucomatous remodeling of the LC likely impact the vascular and mechanical integrity of the optic nerve and may explain the increased vulnerability to further glaucomatous injury seen with aging and with increasing glaucoma severity.

Spectral-domain coherence tomographic imaging (SDOCT) has allowed for the visualization and morphological assessment of the load-bearing components of the ONH, the LC and peripapillary sclera, distinct from the overlying neurovascular tissues they support. This has afforded the opportunity to assess remodeling changes in the LC, which may provide new, mechanistically relevant, biomarkers to predict further glaucomatous injury and identify patients with vulnerable connective tissue architectures. Prior studies have demonstrated age-related and racial variation in the morphology, depth^6-11^, and biomechanical behavior^6^ of the LC, which may explain some of the increased vulnerability to glaucomatous injury seen with aging and in individuals of African Descent.

The African Descent and Glaucoma Evaluation Study (ADAGES) cohort^12^ was developed to evaluate racial differences in the rate, predictors, and mode of progression between individuals of European (ED) and African Descent (AD), given that the AD population has been shown to have a higher prevalence of POAG^13-15^. ADAGES was designed with a methodology that paralleled the ongoing Diagnostic Innovation and Glaucoma Study (DIGS). Imaging data from the combined DIGS/ADAGES cohort has demonstrated that anterior LC surface depth (ALCSD) was more shallow in the ED group, but the ED group exhibited greater posterior migration of ALCSD compared to AD patients^9^. This result suggests that the differences in biomechanical behavior of the LC between AD and ED individuals may result in differences in remodeling responses, which could impact the differential susceptibility to further glaucomatous injury seen across these racial groups.

This study determines the association between visual field (VF) progression rate and depth and curvature of the LC in POAG, followed in the ADAGES and DIGS cohorts in patients using an automated segmentation algorithm. This study will also compare this relationship across AD and ED cohorts to determine if these differences impact the rate of progression and explain some of the increased vulnerability to glaucomatous injury seen in AD populations.

## Methods

This study utilized data obtained from two National Eye Institute-funded longitudinal studies: the University of California at San Diego (UCSD)-based DIGS and the multi-site ADAGES study. DIGS is the longest running structural imaging and functional testing cohort of glaucoma subjects and is conducted only at UCSD. ADAGES (registered at clinicaltrials.gov under NCT00221897) sites include the Hamilton Glaucoma Center at the Department of Ophthalmology, UCSD (data coordinating center), the Edward S. Harkness Eye Institute at Columbia University Irving Medical Center, and the Department of Ophthalmology and Visual Sciences at University of Alabama at Birmingham (UAB). All participants gave written informed consent. The institutional review boards at all three sites approved the study methods. All methods adhered to the tenets of the Declaration of Helsinki and to the Health Insurance Portability and Accountability Act.

Details of the ADAGES and DIGS studies are described previously^9,12^. Both ADAGES and DIGS patients with POAG are followed every 6 months with a comprehensive eye examination, imaging, and automated perimetry. VF testing was performed with a Humphrey perimeter (Carl Zeiss Meditec, Inc.) using the Swedish Interactive Thresholding Algorithm (SITA)-standard 24-2 test pattern. ONH imaging was performed with a Spectralis (OCT2; Heidelberg Engineering GmbH, Heidelberg, Germany). Glaucoma was confirmed at baseline with two reliable repeatable visual field defects consistent with glaucoma as previously described^16^. The UCSD Visual Field Assessment Center reviewed all visual fields for quality throughout the study. The ONH scans used for this analysis were performed with the Spectralis (software version 5.6.4.0 or higher, Heidelberg Engineering, GmbH.) and consisted of 24 radial averaged B-scans. All imaging was evaluated by the Optic Disc and Retinal Reding center and scans with overall poor-quality were excluded as previously described^17^; furthermore, two independent graders (KM and AJ) identified 1,138 (10.8%) of images as unusable for further analysis due to errors in the estimation of the anterior laminar surface by the DL autosegmentation algorithm.

The inclusion and exclusion criteria for ADAGES and DIGS have been previously described^16^. For this study, subject eyes were selected if they had a minimum of 2 years of follow-up and a minimum of 4 exams that included both VF and SDOCT exams. Subjects who received incisional glaucoma surgery were not included. The cohort in ADAGES and DIGS was unbalanced in terms of disease severity and age across racial groups with significantly younger patients with more severe glaucoma in the AD group^17^. To account for this difference, AD and ED subjects were balanced at the baseline-level using Propensity Score Matching. The two Race groups were balanced based on Baseline Age and 24-2 VF MD. In doing so, we controlled for race-related confounding factors in our study.

The analysis for this study was conducted with data from 584 eyes from 359 subjects with POAG (192 ED and 167 AD) that met all SDOCT image quality requirements with eight SDOCT examinations that were propensity matched as described above. Reflectivity (Abyss LLC, Singapore), a commercial deep learning (DL) auto-segmentation software^18^, was used to automatically label the different tissue of the ONH; a custom routine maintained by FastingSoSo (Fast Prototyping Software Solutions LLC; Birmingham, Alabama) written in Wolfram Language (Wolfram Research Inc.; Champaign, IL) estimated the anterior and posterior boundaries on each tissue of interest. ALCSD was estimated in 3D as previously described^19^ using a scleral reference plane based on the 24 radial b-scans. Laminar cribrosa curvature index (LCCI) was calculated as previously described^20^. LCCI is quantified as the average difference between the anterior laminar surface interpolated by a quadratic function^20^ and a reference plane defined by the projections of the BMO points into the anterior lamina surface. This novel approach allows for a definition of a laminar depth metric within the 3D morphology of the lamina itself and standardized against its size (curvature metric) – which disentangles the dependence of the laminar depth from external reference planes and disc size^21,22^

### Statistical analysis

Subject and eye-level demographic and clinical characteristics are presented as count (%) and mean (95% CI) for categorical and continuous parameters, respectively. Subject-level continuous characteristics were evaluated using t-tests for comparisons across AD and ED research groups. Additionally, subject-level categorical characteristics were compared using Fisher’s Exact test. Eye-level continuous characteristics were compared between groups using linear mixed-effects models, fit with random intercepts to account for within-subject variability and intercorrelated measurements. To determine the independent association between longitudinal change in 24-2 SITA MD and baseline ALCSD and LCCI, univariable and multivariable analyses were conducted using linear-mixed effects models, controlling for demographic, clinical and ocular characteristics. Interaction terms were utilized to evaluate the relationships between demographic and clinical characteristics with 24-2 VF MD rate of change (progression rate) over study follow-up time. Linear mixed-effects models which evaluated longitudinal rates of 24-2 VF MD change were fitted with a random intercept to capture between-subject variability at baseline. Measurements of bilateral eyes were nested within subject to account for correlated measurements from eyes belonging to the same subject. All statistical analyses were performed using the R programming language for statistical computation (R Core Team (2024). R Foundation for Statistical Computing, Vienna, Austria.). Measurements of bilateral eyes were nested within subject to account for correlated measurements from eyes belonging to the same subject. These linear mixed-effects models also incorporated autocorrelation structures (corExp function; NLME package) to address temporal dependence across longitudinal repeated measures, and fixed variance weights to address within-subject heteroscedasticity. P-values less than 0.05 were considered statistically significant.

## Results

Demographic and clinical characteristics of the subjects and eyes are shown in Table 1. After matching for baseline age and 24-2 VF MD we included 292 eyes (167 participants) and 292 eyes (192 participants) of AD and ED, respectively. The AD patients were younger than the ED patients (62.6 vs 64.8) but did not reach statistical significance (p=0.056). The average mean deviation in VF over the follow-up was -3.42 dB for the AD group and -2.56 dB for ED group (p=0.091). AD patients had thinner central corneal thickness (CCT) (535.6 µm vs 551.6 µm; p<0.001). Axial length, IOP and LCCI were similar across groups. ALCSD was deeper in the AD than the ED group (385 microns vs. 363 microns) but this did not reach statistical significance (p=0.052).

**Table 1.** Demographic and Ocular Characteristics across racial groups propensity matched for age and visual field severity.

|  | African Descent<br>(n = 167 subjects;<br>292 eyes) | European Descent<br>(n = 192 subjects; 292<br>eyes) | Overall<br>(n = 359 subjects; 584<br>eyes) | p-value |
| --- | --- | --- | --- | --- |
| Age (years) | 62.6 (60.7, 64.4) | 64.8 (63.3, 66.3) | 63.8 (62.6, 64.9) | 0.056 |
| Sex |  |  |  |  |
| Female | 110 (65.9%) | 110 (57.3%) | 220 (61.3%) | 0.104 |
| Male | 57 (34.1%) | 82 (42.7%) | 139 (38.7%) |  |
| Mean 24-2 VF MD across follow-up<br>(dB) | -3.42 (-4.14, -2.70) | -2.56 (-3.25, -1.87) | -2.97 (-3.47, -2.47) | 0.091 |
| 24-2 VF MD at baseline (dB) | -3.20 (-3.89, -2.52) | -2.23 (-2.89, -1.58) | -2.70 (-3.17, -2.22) | **0.046** |
| Axial Length (mm) | 24.0 (23.8, 24.1) | 24.0 (23.8, 24.1) | 24.0 (23.8, 24.1) | 0.995 |
| IOP (mmHg) | 15.47 (14.88, 16.06) | 15.78 (15.22, 16.34) | 15.63 (15.23, 16.04) | 0.453 |
| CCT (μm) | 535.6 (529.5, 541.8) | 551.6 (545.8, 557.4) | 544.2 (539.9, 548.5) | **< 0.001** |
| Lamina Depth (μm) | 385.2 (368.7, 401.7) | 362.9 (347.6, 378.1) | 373.2 (361.9, 384.4) | 0.052 |
| LCCI (Curvature) | 0.12 (0.12, 0.13) | 0.12 (0.11, 0.13) | 0.12 (0.12, 0.13) | 0.465 |
| VF MD = visual field mean deviation; IOP = intraocular pressure; CCT = central corneal thickness; LCCI = lamina cribrosa curvature index |  |  |  |  |

Table 2 summarizes the results from univariable, full multivariable, and final models for the analyses evaluating the ability of ALCSD to predicted longitudinal decay in visual field mean defect (MD) across follow up time. These linear mixed effects models report the variable of interest as a main effect plus their interaction term with time. The final model was chosen from the clinically relevant parameters measured based on minimization of Conditional Akaike information criterion index (AICc). In the full multivariable model and the final model, ALCSD, race, IOP and visual field MD were all independently associated with visual field progression. In the final model, a deeper ALCSD (−0.02 dB/year per 50 microns, p<0.001), advancing age (−0.025dB/year per 10 years, p-0.007), European descent (−0.056 dB/year, p=0.006), higher IOP (− 0.011dB/year per 2mmHg, p=0.021), and more severe visual field defect (0.017dB/year per 1dB, p<0.001) were all independently associated with a faster progression rate.

**Table 2.** Linear Mixed-Effects Model Results for the Relationship Between Anterior Lamina Cribrosa Surface Depth and Change in Visual Field Mean Defect.

|  | Univariable Results |  |  | Full Multivariable Results |  |  | Final Multivariable Model |  |  |
| --- | --- | --- | --- | --- | --- | --- | --- | --- | --- |
| Variable | Coefficient Estimate (95% CI) | Standard Error | p-value | Coefficient Estimate (95% CI) | Standard Error | p-value | Coefficient Estimate (95% CI) | Standard Error | p-value |
| **Predictors** |  |  |  |  |  |  |  |  |  |
| **Intercept** |  |  |  | -2.385<br>(-2.466, -2.304) | 0.041 | < **0.001** | -2.371<br>(-2.450, -2.291) | 0.04 | < **0.001** |
| Follow-Up Time (years) | -0.189<br>(-0.217, -0.162) | 0.014 | < **0.001** | -0.073<br>(-0.102, -0.043) | 0.015 | < **0.001** | -0.075<br>(-0.103, -0.047) | 0.014 | < **0.001** |
| Baseline Lamina Depth, per 50µm deeper | -0.677<br>(-0.908, -0.446) | 0.117 | < **0.001** | 0.046<br>(0.021, 0.072) | 0.013 | < **0.001** | 0.044<br>(0.019, 0.070) | 0.013 | < **0.001** |
| Baseline Age, per 10 years older | -0.392<br>(-0.837, 0.052) | 0.226 | 0.083 | 0.058<br>(0.008, 0.108) | 0.025 | **0.024** | 0.066<br>(0.017, 0.115) | 0.025 | **0.009** |
| Race, European Descent | 0.833<br>(-0.172, 1.837) | 0.511 | 0.104 | 0.063<br>(-0.045, 0.171) | 0.055 | 0.249 | 0.053<br>(-0.050, 0.157) | 0.053 | 0.312 |
| IOP, per 2 mmHg higher | 0.078<br>(0.028, 0.128) | 0.025 | **0.002** | 0.030<br>(0.001, 0.060) | 0.015 | **0.043** | 0.021<br>(-0.006, 0.049) | 0.014 | 0.127 |
| CCT, per 20µm thicker | 0.040<br>(-0.058, 0.139) | 0.05 | 0.422 | 0.004<br>(-0.022, 0.030) | 0.013 | 0.768 |  |  |  |
| Mean 24-2 VF MD across Follow-up (dB) | 0.955<br>(0.943, 0.967) | 0.006 | < **0.001** | 0.917<br>(0.896, 0.939) | 0.011 | < **0.001** | 0.919<br>(0.898, 0.941) | 0.011 | < **0.001** |
| **Interaction Terms** |  |  |  |  |  |  |  |  |  |
| Follow-Up Time * Baseline Lamina Depth, per 50µm deeper | -0.026<br>(-0.038, -0.014) | 0.006 | < **0.001** | -0.021<br>(-0.030, -0.011) | 0.005 | < **0.001** | -0.020<br>(-0.029, -0.011) | 0.005 | < **0.001** |
| Follow-Up Time * Baseline Age, per 10 years older | -0.059<br>(-0.084, -0.035) | 0.012 | < **0.001** | -0.020<br>(-0.040, -0.000) | 0.01 | **0.047** | -0.025<br>(-0.043, -0.007) | 0.009 | **0.007** |
| Follow-Up Time * Race, European Descent | -0.005<br>(-0.060, 0.051) | 0.028 | 0.865 | -0.070<br>(-0.113, -0.027) | 0.022 | **0.001** | -0.056<br>(-0.096, -0.016) | 0.020 | **0.006** |
| Follow-Up Time * IOP, per 2 mmHg higher | -0.043<br>(-0.056, -0.031) | 0.006 | < **0.001** | -0.020<br>(-0.032, -0.009) | 0.006 | < **0.001** | -0.011<br>(-0.021, -0.002) | 0.005 | **0.021** |
| Follow-Up Time * CCT, per 20µm thicker | 0.003<br>(-0.011, 0.017) | 0.007 | 0.710 | 0.003<br>(-0.007, 0.013) | 0.005 | 0.618 |  |  |  |
| Follow-Up Time * Mean 24-2 VF MD across Follow-up (dB) | 0.023<br>(0.018, 0.028) | 0.002 | < **0.001** | 0.020<br>(0.011, 0.028) | 0.004 | < **0.001** | 0.017<br>(0.010, 0.025) | 0.004 | < **0.001** |
| VF MD = visual field mean deviation; IOP = intraocular pressure; CCT = central corneal thickness; LCCI = lamina cribrosa curvature index |  |  |  |  |  |  |  |  |  |

The univariable associations with visual field progressin for demographic and ocular characteristics along with baseline LCCI are shown in Table 3 along with the fill and final models. Similar to ALCSD, the rate of progression was significantly increased with increasing age and IOP, with more severe visual fields, and was faster in the ED group in both full and final models. In the final model, greater LC posterior curvature (higher LCCI, -0.01dB/year per 0.01units, p<0.001), advancing age (−0.023 dB/year per 10 years, p=0.013), European descent (−0.063 dB/year, p=0.001), higher IOP (−0.019 dB/year per 2mmHg, p<0.001), and more severe visual field defect (0.034dB/year per 1dB, p<0.001) independently predicted more rapid visual field progression.

**Table 3.**
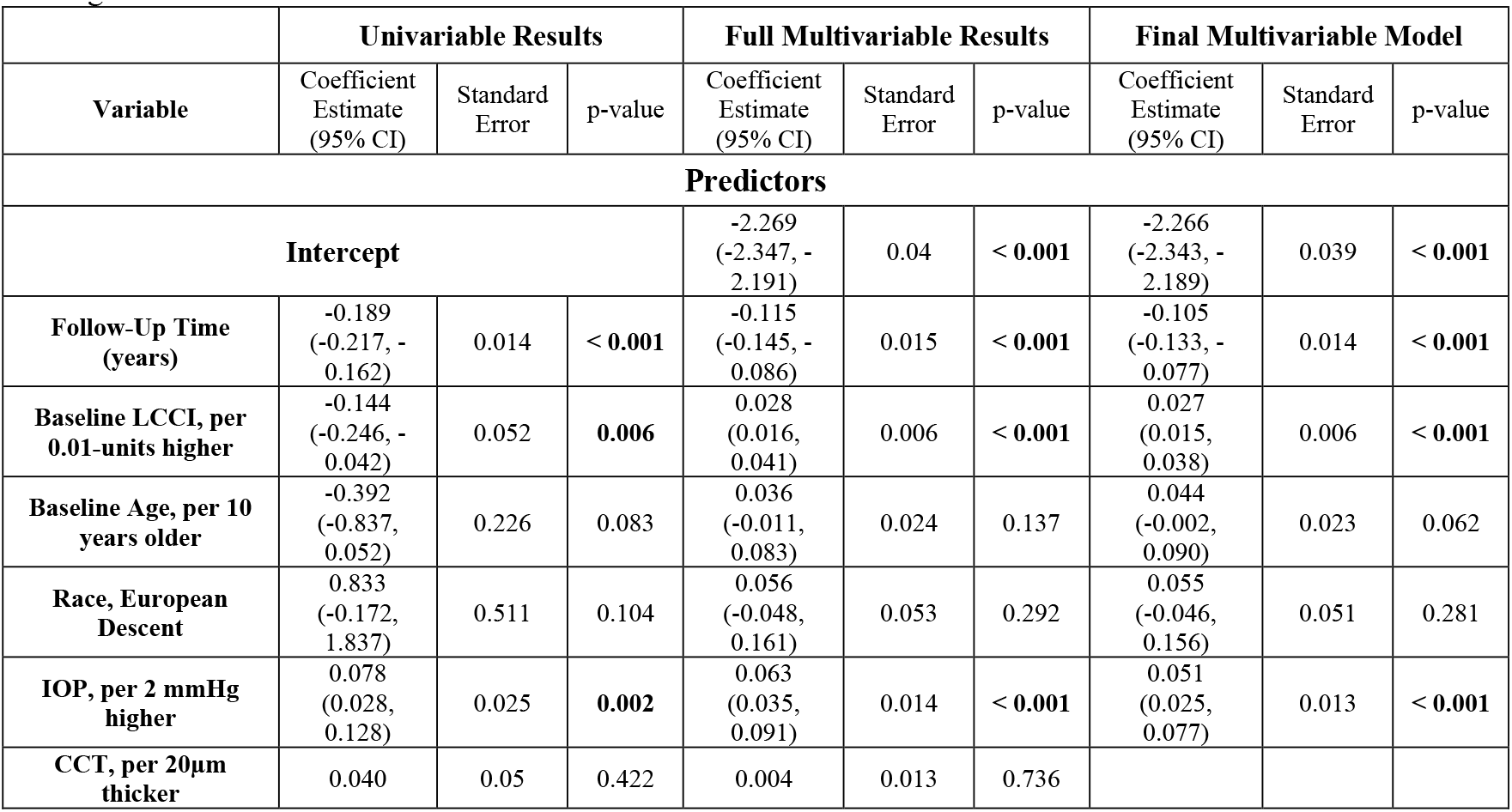
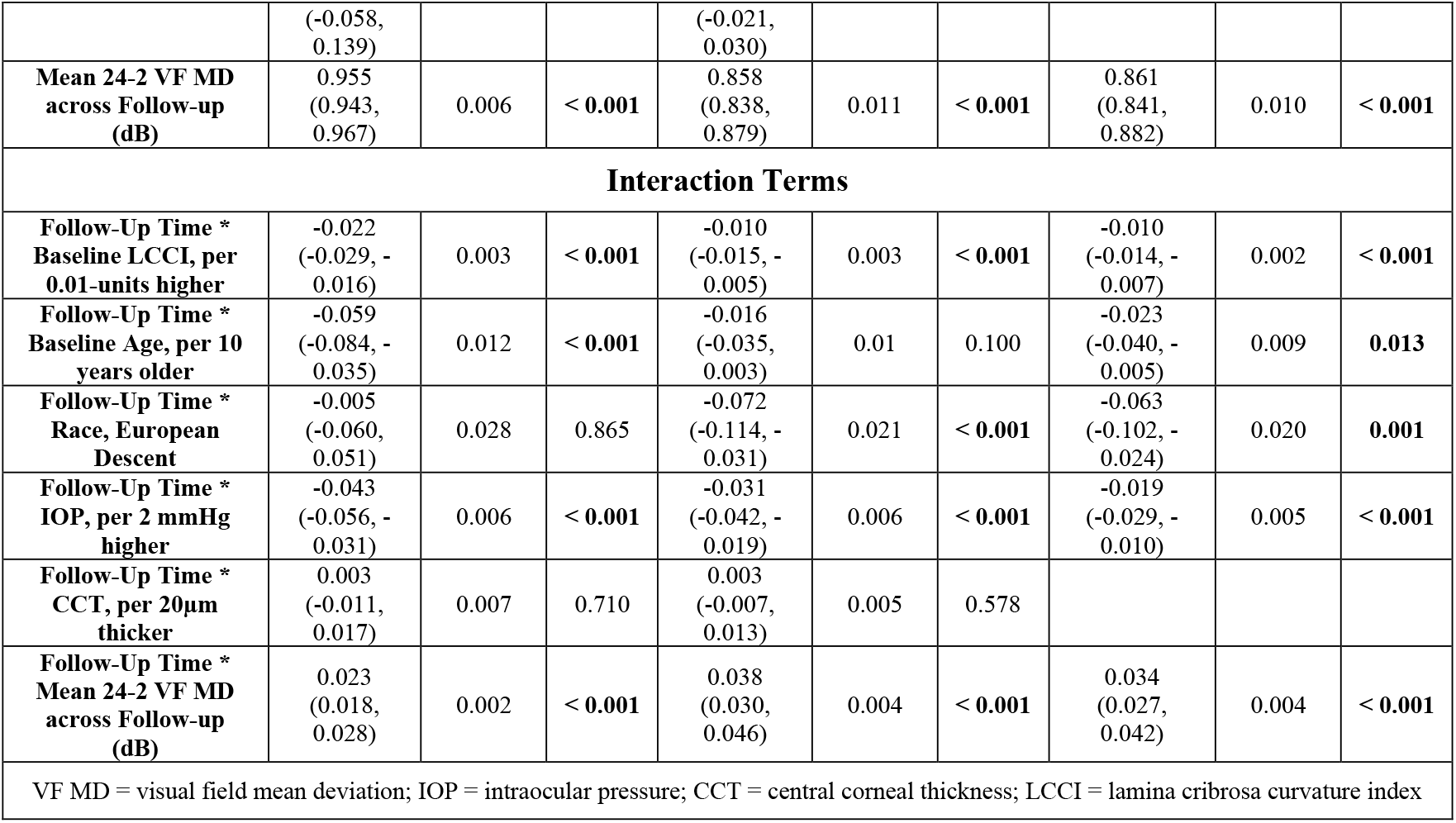
Linear Mixed-Effects Model Results for the Relationship Between Lamina Cribrosa Curvature Index and Change in Visual Field Mean Defect.

Figure 1 and 2 illustrates the change in linear relationships between the decline in visual field MD over time across quartiles and mean estimates for each of the continuous variables that remained significant within the final multivariable model for ALCSD and LCCI respectively. The magnitude of change across these quartiles provided some comparison of the relative impact of the effect of the variation in these individual parameters on the rate progressive visual field loss, while accounting for differences in the range, units and distributions of the individual continuous parameters. All other continuous predictors within the final multivariable models were set to their mean value, while categorical predictors were set to their reference level.

**Figure 1:**
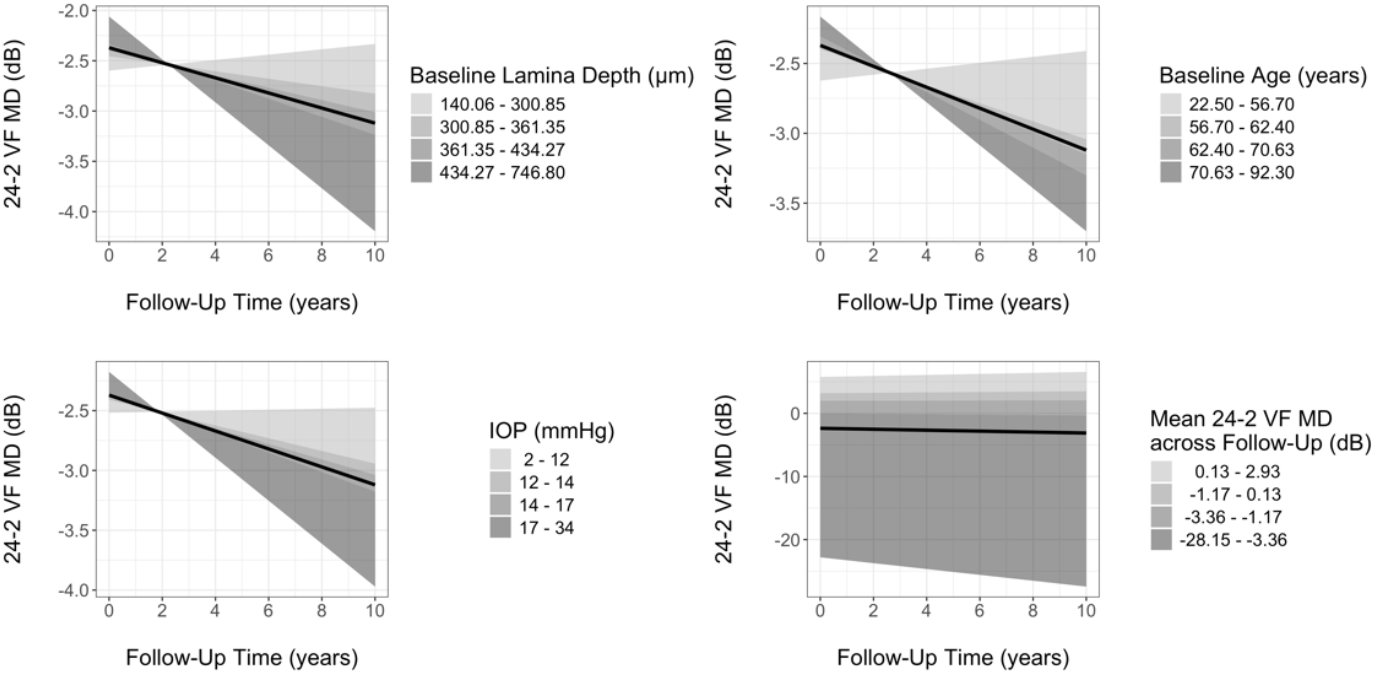
Linear Relationship between the rate of visual field progression (24-2 VF MD) across ordinal quartiles of measurements of baseline anterior lamina cribrosa surface depth, age, IOP and baseline visual field severity. Black line represented the overall mean.

**Figure 2.**
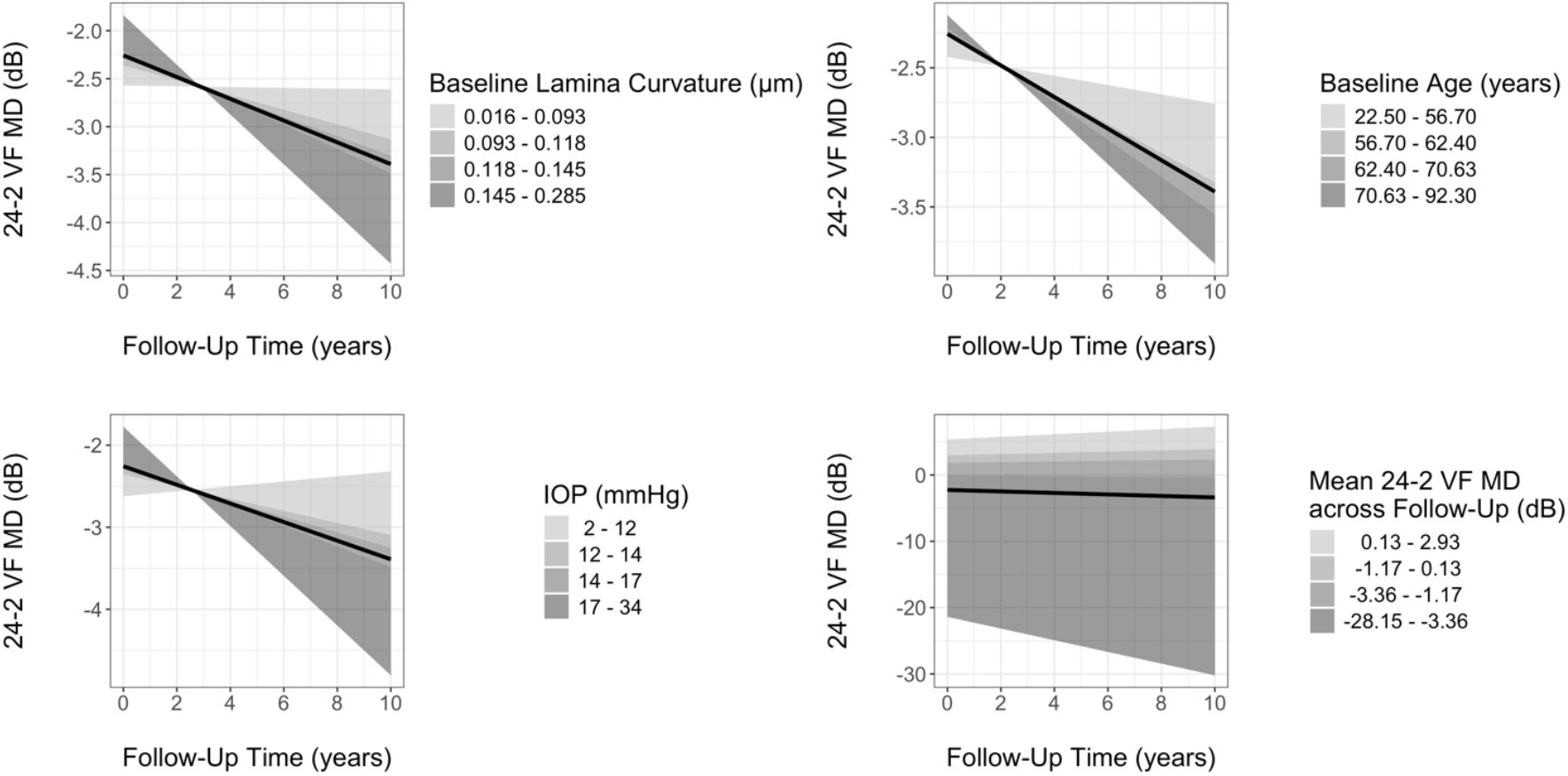
Linear Relationship across the rate of visual field progression (24-2 VF MD) across ordinal quartiles of measurements of baseline lamina cribrosa curvature index, age, IOP and baseline visual field severity. Black line represented the overall mean.

Figure 3 illustrates the change in the slope of visual field loss for AD and ED groups from the adjusted analyses for ALCSD (Figure 3a) and LCCI (Figure 3b). The AD group demonstrated slower rates of decline in SITA 24-2 VF MD in both models when compared to the ED group.

**Figure 3.**
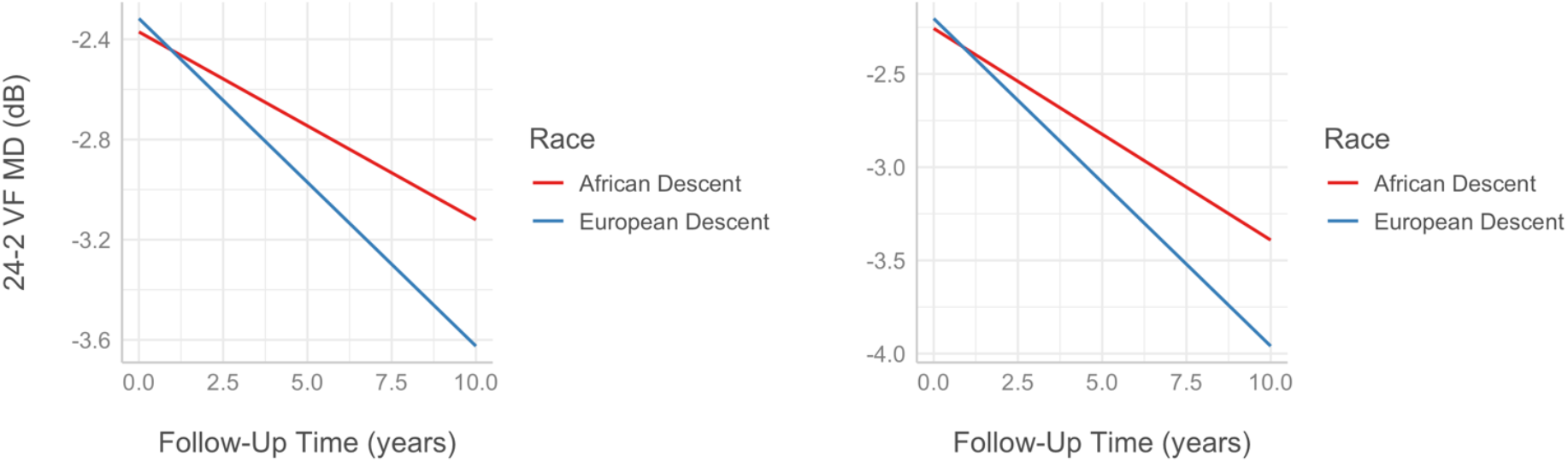
Linear rate of decline in visual field mean defect (24-2 VF MD) for participants of African and European Descent for the multivariable model using anterior lamina cribrosa surface depth (**left**) and lamina cribrosa curvature index (**right**).

## Discussion

This study utilized longitudinal data from the ADAGES and DIGS cohort studies to determine the association between baseline lamina cribrosa depth and curvature, their interactions with self-described race, and VF progression. Our findings suggest that deeper laminar depth and more curved laminar surface are independently associated with faster VF progression. The magnitude of impact on the rate of visual field decay was similar to the impact seen with IOP and aging within these variables’ ranges and distributions in the dataset (Figure 1 and 2). This finding aligns with the hypothesized importance of LC morphology to the mechanovascular behavior of the ONH that may ultimately impact the vulnerability of the optic nerve to glaucomatous injury^5,23,24^.

Remodeling of the load bearing connective tissues of the LC is a critical histopathologic characteristic of glaucomatous optic neuropathy^25^. Pathologic and age-related remodeling of the LC may result in alterations in the morphology and material properties of these supportive load bearing tissues and is hypothesized to increase the risk of further glaucomatous injury. Changes in the morphology of the LC has been investigated in non-human primate models^26-33^, human donor tissues^23,34-36^, and in vivo imaging studies^37-39^. In humans and primates, in vivo imaging studies have indicated that the ALCSD may vary depending on the disease stage and can potentially move to a more anterior or posterior location within the scleral canal^9,10,22,39,40^.

While several cross-sectional studies have examined the role of ALCSD in glaucoma patients^10,40-42^, to our knowledge only one previous cross-sectional study has examined the relationship between ALCSD and glaucoma severity in individuals of African and European Descent, with known differences in optic nerve morphology and biomechanics^22^ and differential susceptibility to glaucomatous injury. This prior cross-sectional study also utilized data from the ADAGES and DIGS cohort and evaluated racial differences in the relationship between ALCSD, disease severity, and ocular and demographic parameters in glaucomatous eyes of AD and ED. Overall, a deeper ALCSD was independently associated with higher IOP, thicker peripapillary choroid, male gender, African descent, and a larger optic nerve area. Conversely, a shallower ALCSD was associated with advancing age and an increased visual field mean defect. The AD group exhibited shallower ALCSD associated with increasing age and higher IOP. These cross-sectional findings demonstrated that factors associated with variations in the morphometry of the deep optic nerve head in glaucomatous optic neuropathy may differ across racial groups and may interact with aging and glaucoma related remodeling of these load bearing connective tissues. This suggests that age-related and glaucomatous remodeling of the optic nerve head may differ between these racial groups.

There have been few longitudinal studies examining the relationship of ALCSD with aging and glaucoma^9,39,43-45^. While previous cross-sectional studies indicated that increased glaucoma severity was associated with a deeper LC, these longitudinal studies have shown that remodeling of the LC results in both anterior and posterior displacement of ALCSD over time. We previously conducted a longitudinal study of ALCSD change over time using data from the ADAGES and DIGS cohorts that demonstrated racial disparities in the rate and mode of progressive changes in LC depth among glaucoma patients^9^. This previous study demonstrated that patients of ED showed more pronounced posterior migration of the LC compared to patients of AD. These differences were observed independently of racial variations in optic disc size, as measured by the Bruch’s membrane opening area.

Several cross-sectional studies have examined the relationship between LCCI and glaucoma, demonstrating that LCCI changes with acute changes in IOP^46^, can discriminate between normal and glaucomatous eye^20^, and between eyes with ocular hypertension and normal tension glaucoma^47^. There was also a higher discriminatory value for LCCI when compared to ALCSD^20^. In correspondence to our findings, longitudinal studies have demonstrated increasing LCCI is associated with progressive nerve fiber layer thinning in glaucoma patients^48^ and glaucoma suspects^44^, and predicts more rapid visual field decline^45^. In the current study, LCCI was similar across AD and ED groups. This study also confirms that greater posterior curvature of the LC in the ADAGES cohort, as reflected in a high LCCI, is an independent structural biomarker for glaucoma. While covariance would not allow for the direct comparison of LCCI and ALCSD within the same model, the magnitude of effect based on parameter estimates (tables 1 and 2, Figure 1 and 2) appears similar for LCCI and ALCSD.

In the current study, while the laminar surface was deeper in the AD group, the AD group demonstrated a slower rate of visual field progression. This unexpected finding was first observed in a prior study utilizing ADAGES/DIGS data^17^. However, the overall ADAGES/DIGS cohort, due to being a convenient sample obtained from tertiary referral clinics, was imbalanced across racial groups, with a significantly lower age and significantly more severe glaucoma in the AD group. This current study utilized propensity matching across age and disease severity to account for this imbalance in comparing the progression rate across racial groups and the interactions of self-described race with the relevant associated covariables. Even with this rebalancing, the ED group progressed faster than the AD group. These finding seem to conflict with population-based data and other cohort studies which show the populations of sub-Saharan African descent developed glaucoma earlier, progression more rapidly and are at greater risk of blindness from glaucoma^49,50^. One possible explanation is that ADAGES and DIGS provided medications, patient payments, and transportation to participants and also utilized clinical coordinators trained in cultural competency^12^. These factors may have addressed several barriers that may impair healthcare engagement. This suggests that these socio-economic and cultural barriers play a far greater role in the greater burden of glaucoma in minoritized populations than any intrinsic predisposition to develop and progress from glaucoma.

The study has a several limitations. While patients that received surgical glaucoma therapy were censored, all patients were under medical treatment for glaucoma and thus may not reflect the effects seen in an untreated population and may confound the association between IOP and progression. The participants overall had relatively mild glaucoma based on VF MD. Therefore, these results may not be applicable to patients with more advanced glaucoma. Moreover, all imaging was performed by highly trained research personnel which may impact image quality and generalizability for the use of ALCSD and LCCI as biomarkers. However, the analytic approaches used in this study are readily deployable to existing clinical devices.

In summary, this study demonstrated that a deeper, more posteriorly curved, laminar surface was independently associated with visual field progression in patients with POAG. This study also confirms in a balanced cohort that the ED group progressed faster than the AD group within the ADAGES cohort. These results support that the hypothesis that alterations in laminar cribrosa morphology due to glaucomatous remodeling of the load bearing connective tissues of the optic nerve head increased the vulnerability to more rapid functional progression.

## Data Availability

All data produced in the present study are available upon reasonable request to the authors

## Abbreviations

AD: African Descent
ADAGES: African Descent and Glaucoma Evaluation Study
AICc: Akaike information criterion index
ALCSD: Anterior Lamina Cribrosa Surface Depth
CTT: Central Corneal Thickness
DIGS: Diagnostic Innovation and Glaucoma Study
DL: Deep Learning
ED: European Descent
IOP: Intraocular Pressure
LC: Lamina Cribrosa
LCCI: Laminar Cribrosa Curvature Index
ONH: Optic Nerve Head
POAG: Primary Open Angle Glaucoma
RGC: Retinal ganglion cells
SDOCT: Spectral Domain Optic Coherence Tomography
SITA: Swedish Interactive Thresholding Algorithm
VF: Visual Field
VF MD: Visual Field Mean Deviation
UAB: University of Alabama-Birmingham
UCSD: University of California, San Diego
US: United States

